# TAME-Q: An Open-Source Preprocessing Pipeline for Reproducible Semi-Quantification of Florzolotau (18F) PET

**DOI:** 10.1101/2025.10.08.25337562

**Authors:** Kenjiro Nakayama, Kiyotaka Nemoto, Hironobu Endo, Kenji Tagai, Asaka Oyama, Miyuki Nemoto, Masashi Tamura, Miho Ota, Takahiko Tokuda, Makoto Higuchi, Tetsuaki Arai

**Author notes:** Corresponding Author: Kiyotaka Nemoto, Department of Medical Informatics and Management and Psychiatry, Institute of Medicine, University of Tsukuba, Address: 1-1-1 Tennodai Tsukuba, Ibaraki, 305-8575, Japan.

## Abstract

**Purpose:** Accurate semi-quantification of florzolotau (18F) PET in Alzheimer’s disease (AD) and non-AD tauopathies is essential. While histogram-based approaches are promising, their implementation relies on proprietary software, limiting accessibility. Thus, we developed TAME-Q (Tomographic Image Preprocessing Using Automated Multi-Reference Estimation for Quantification), an open-source preprocessing pipeline that performs histogram-based semi-quantification. This study aimed to use this open-source framework to produce results consistent with those of prior work and to validate the pipeline’s performance for florzolotau PET semi-quantification.

**Methods:** This study included 37 individuals on the AD continuum, 46 with probable progressive supranuclear palsy-Richardson syndrome (PSP-RS), and 50 healthy controls. TAME-Q was used to co-register T1-weighted MR and florzolotau PET images and perform semi-quantification by fitting a bimodal Gaussian mixture to grey-matter signal intensities. Elastic Net models yielded AD-tau and PSP-tau scores indicating disease-characteristic tau burdens. The standardized uptake value ratios (SUVRs) of the inferior temporal gyrus (ITG) and globus pallidus (GP) were compared between groups. The two pipelines were compared via correlation analysis and ROC/AUC comparison using DeLong’s test.

**Results:** TAME-Q processed all subject data without manual correction. The SUVR was elevated specifically in the ITG for AD and the GP for PSP. Tau scores correlated with previous scores (AD-tau: R = 0.983; PSP-tau: R = 0.966) and had AUCs of 0.999 (AD) and 0.977 (PSP-RS) with no significant difference in discriminative performance.

**Conclusion:** TAME-Q enables fully automated and reproducible semi-quantification of florzolotau PET comparable to previous methods. Its open-source implementation supports transparent dissemination and broader adoption in research settings.

## 1 Introduction

Positron emission tomography (PET) has revolutionized the study of neurodegenerative diseases by enabling in vivo visualization of misfolded protein aggregates, including amyloid plaques [1–6], tau aggregates [7–14], and α-synuclein aggregates [15]. In Alzheimer’s disease (AD), amyloid and tau PET imaging have reshaped diagnostic criteria, allowed precise staging of disease progression, and provided indispensable biomarkers in clinical trials for patient selection and monitoring of therapeutic efficacy [16–18].

Despite these advances in AD research, accurate diagnosis of non-AD tauopathies—most notably progressive supranuclear palsy (PSP), corticobasal degeneration (CBD), and frontotemporal lobar degeneration—remains challenging due to clinical heterogeneity and symptom overlap with other parkinsonian syndromes. Although tau PET could overcome these challenges, most first- and second-generation tau tracers— particularly flortaucipir (18F) analogs (e.g., [^18^F]RO948) and [^18^F]MK-6240—do not exhibit a high affinity for non-AD tau aggregates [19–22].

In contrast, the tau PET tracer florzolotau (18F) ([^18^F]PM-PBB3/[^18^F]APN-1607) overcomes this limitation. It binds both AD and non-AD tau aggregates with high affinity in human postmortem tissue, and early in vivo studies suggest that florzolotau captures distinct regional deposition patterns in PSP, corticobasal syndrome, and Pick’s disease [12,23]. To convert these qualitative observations into robust quantitative biomarkers, semi-quantitative analysis methods are required to standardize measurements across individuals, support longitudinal tracking, and integrate with machine learning classifiers. A fundamental challenge is the selection of an appropriate reference region for calculating standardized uptake value ratios (SUVRs). Although the cerebellar cortex is used as a reference region in AD studies due to its relative sparing from tau pathology [24], it is theoretically inappropriate for PSP and related disorders in which tau deposition can extend to the cerebellum [25]. Therefore, it is imperative to identify an optimal reference region that can accommodate these complexities.

A histogram-based, data-driven approach has been proposed to address this issue. This method fits a bimodal distribution to voxel intensities in grey matter and selects the lower intensity mode as the reference [26], thereby avoiding regions with subtle pathology and offering an unbiased alternative to conventional references. Combined with machine learning applied to ROI (region of interest)-based SUVRs, this method has been shown to identify key regions for differential diagnosis and yield composite tau scores with high diagnostic accuracy that correlates with clinical severity [27]. However, the utility of this advanced method is constrained by its reliance on in-house or proprietary software, which limits reproducibility and broader adoption.

To overcome this barrier, we have developed TAME-Q (Tomographic Image Preprocessing Using Automated Multi-Reference Estimation for Quantification), a fully automated, open-source preprocessing pipeline. The primary aim of this study is, therefore, twofold. First, we seek to produce results that are consistent with a previous benchmark study [27] using the entirely of our open-source framework. Second, through this process, we aim to validate TAME-Q by demonstrating that it processes florzolotau PET data accurately without manual intervention and achieves a diagnostic performance comparable to the current standard method. By establishing the reproducibility and validity of this automated pipeline, we aim to standardize florzolotau PET analysis, accelerate biomarker discovery, and facilitate future therapeutic trials targeting tauopathies.

## 2 Materials and methods

### 2.1 Participants

This study used the dataset described in an earlier publication [27], which comprised 37 participants on the AD continuum, 46 individuals with the probable progressive supranuclear palsy-Richardson syndrome (PSP-RS), and 50 healthy controls (HC). AD continuum diagnoses were based on Petersen’s criteria [28] for mild cognitive impairment and the criteria of the National Institute of Neurological and Communicative Diseases and Stroke/Alzheimer’s Disease and Related Disorders Association [29]. PSP-RS was diagnosed following the International Parkinson and Movement Disorder Society clinical diagnostic criteria for PSP [30]. Healthy controls satisfied the following criteria: aged ≥40 years, no history of neurological diseases or depression, visually negative on [^11^C]Pittsburgh Compound-B (PiB) PET, Mini-Mental State Examination (MMSE) score ≥28 or Montreal Cognitive Assessment (MoCA) score ≥26, and Geriatric Depression Scale (GDS) score ≤5.

Written informed consent was provided from every participant. For participants with cognitive impairments, written informed consent was additionally obtained from their spouses or close relatives as appropriate. This study was approved by the Radiation Drug Safety Committee and the National Institutes for Quantum Science and Technology Certified Review Board of Japan, and the Ethics Committee of the University of Tsukuba Hospital in accordance with the Declaration of Helsinki (R02-328).

### 2.2 Image acquisition

All participants underwent three-dimensional T1-weighted MR and florzolotau PET scans. MR scans were acquired on a 3-T MAGNETOM Verio (Siemens) scanner in sagittal orientation, with a repetition time of 2,300 ms, echo time of 1.95 ms, flip angle of 9°, slice thickness of 1.0 mm, and matrix size of 512 × 512 × 176.

PET imaging commenced 90-110 minutes after intravenous injection of 186 ± 8.4 MBq florzolotau and was performed on a Biograph mCT flow system (Siemens Healthcare) using a voxel size of 2 × 2 × 2 mm. Data were acquired in either two 10-minute frames or four 5-minute frames and reconstructed by filtered back-projection with a 6.0 mm full width at half maximum Hanning filter.

### 2.3 TAME-Q pipeline implementation

The overall workflow of the TAME-Q pipeline is shown in Fig. 1a. The TAME-Q pipeline integrates FMRIB Software Library (FSL) [31,32], Statistical Parametric Mapping 12 (SPM12; Welcome Trust Centre for Neuroimaging, UCL, UK), FreeSurfer [33–35], and Python 3 with several libraries including NiBabel and SciPy [36] to process florzolotau PET and T1-weighted MRI data from alignment through semi-quantification. Inputs and outputs are in Neuroimaging Informatics Technology Initiative (NIfTI) format, with final outputs comprising SUVR images and ROI-level SUVR tables. These analyses were executed on Lin4Neuro (Ubuntu 22.04-based Linux distribution) [37].

**Fig. 1.**
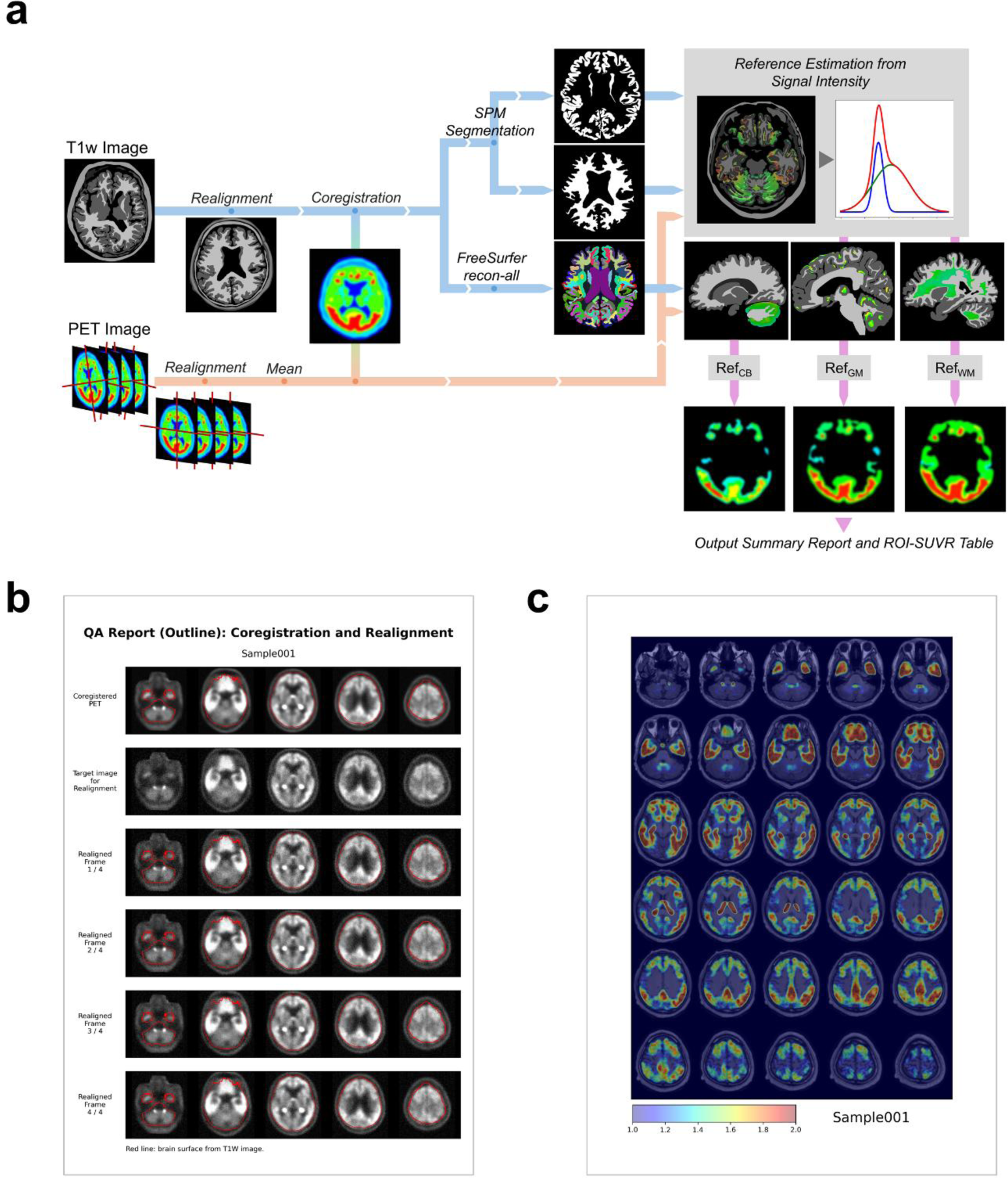
(a) Flowchart of the preprocessing pipeline. First, the T1-weighted image and PET image are realigned and co-registered. Then, the T1-weighted image is segmented into regions. Signals in grey matter are used to calculate reference values based on the histogram method (described in previous research). Finally, the PET image is semi-quantified by the reference value. The images shown here are for illustrative purposes and do not represent actual data. (b) A report image for quality assurance. Proper realignment and co-registration can be verified by confirming whether the averaged PET image and each individual PET frame align with the outer contour of the T1-weighted image. (c) An overview image is exported in a format that does not require specialized neuroimaging software, enabling easy visual inspection of the processed data.

Initially, skull stripping was performed on the T1-weighted images using SynthStrip [38]. The brain images were then rigidly aligned to the Montreal Neurological Institute (MNI) brain template [39] using FLIRT (FMRIB’s Linear Image Registration Tool) [40]. Dynamic PET frames were realigned to the first frame, averaged into a single static scan, and co-registered to the T1 image. To confirm proper alignment, a quality assurance (QA) report was generated. The report overlays the T1-derived brain surface on each PET frame as shown in Fig 1b.

For tissue segmentation, SPM12 was used to generate probability maps of grey matter, white matter, and cerebrospinal fluid (CSF). To minimize signal contamination from surrounding tissues, erosion was performed on the grey and white matter masks using a kernel that incorporates four adjacent voxels in the x- and y-axis directions for any given voxel.

To determine reference values, PET signals within the grey matter mask were binned in 0.025-unit increments and modeled as the sum of two Gaussians fitted via Levenberg-Marquardt algorithm in SciPy library 1.15.1 [36]. The Gaussian component with the lower mean, corresponding to non-tau-accumulated regions, was identified. Voxels in the grey matter region with signal values within the lower curve’s full width at half maximum (FWHM) were defined as the reference region, and their mean intensity was designated as the reference value. If the Gaussian curve with the smaller mean had a peak height less than half that of the curve with the larger mean, additional monomodal curve fitting was performed. When the Dice coefficient between the monomodal distribution and the target distribution was ≥ 0.936, the outcome of the monomodal fitting was adopted. In such cases, voxels with signal intensities within FWHM of the monomodal curve were used to define the reference region, and their mean intensity served as the reference value.

SUVR images were then generated by dividing PET intensities by the reference value. These SUVR images were also output as lightbox views to facilitate visual assessment as shown in Fig 1c. To define ROIs, T1-weighted images were segmented into 162 regions using the recon-all pipeline implemented in FreeSurfer 7.4.1 [33–35].

Brainstem structures were further subdivided using the automated Bayesian segmentation package [41]. To harmonize with the parcellation scheme used in previous studies [27], specified ROIs (Table 1) were merged, yielding a final set of 133 ROIs.

**Table 1.** Merged regions from original FreeSurfer output to the proposed atlas.

| Original Regions | Merged Region |
| --- | --- |
| caudal middle frontal, rostral middle frontal | middle frontal |
| pars opercularis, pars triangularis, pars orbitalis | inferior frontal |
| lateral orbitofrontal, medial orbitofrontal, frontal pole | orbitofrontal |
| caudal anterior cingulate, isthmus cingulate, posterior cingulate, rostral anterior cingulate | cingulate |

All intermediate outputs, including the QA reports and the lightbox views of SUVR images, underwent visual quality control to confirm successful realignment, segmentation, and curve fitting.

### 2.4 Machine learning analysis for detecting disease-specific tau distribution

To validate TAME-Q’s preprocessing and ROI selection, we derived tau scores from TAME-Q outputs. Following the method used in a previous study [27], we trained Elastic Net logistic regression models using age- and sex-adjusted (z-scored) ROI SUVRs to distinguish AD from the combined HC+PSP group, and PSP from the combined HC+AD groups. We partitioned the dataset into 75% training and 25% validation sets via stratified shuffle-split in scikit-learn, preserving sex ratios across splits. We repeated this split 10 times and, within each training set, conducted 5-fold cross-validation to tune the Elastic Net’s *L1_ratio* and *α* parameters, optimizing the area under the receiver operating characteristic curve (ROC/AUC). After cross-validation, we averaged the optimal hyperparameters across folds and repeats, then averaged the resulting model coefficients to acquire the final equations for the AD-tau and PSP tau scores.

### 2.5 Statistical analysis

We assessed demographic differences by one-way analysis of variance (ANOVA) across the HC, AD, and PSP groups, and assessed sex distribution using a chi-squared test. To evaluate the effectiveness of the proposed method using TAME-Q, we compared SUVRs in predefined disease-specific ROIs: the inferior temporal gyrus (ITG) for AD and the globus pallidus (GP) for PSP. For these ROIs, we averaged the left- and right-ROI SUVRs, then performed Welch’s t-test between groups.

Next, to verify comparability with existing methods, we computed Pearson correlation coefficients between tau scores derived from TAME-Q and those from the benchmark method [27], which utilized M-Vision (M Corporation, Tokyo, Japan) for structural parcellation and in-house scripts for standardization. We then evaluated discriminative performance by plotting ROC curves for the AD-tau score (AD vs. HC+PSP) and the PSP-tau score (PSP vs. HC+AD) and compared AUCs using DeLong’s test between the benchmark and proposed methods. All statistical analyses were performed using R version 4.4.2 (R Core Team, 2024).

## 3 Results

Participant demographics are summarized in Table 2.

**Table 2.**
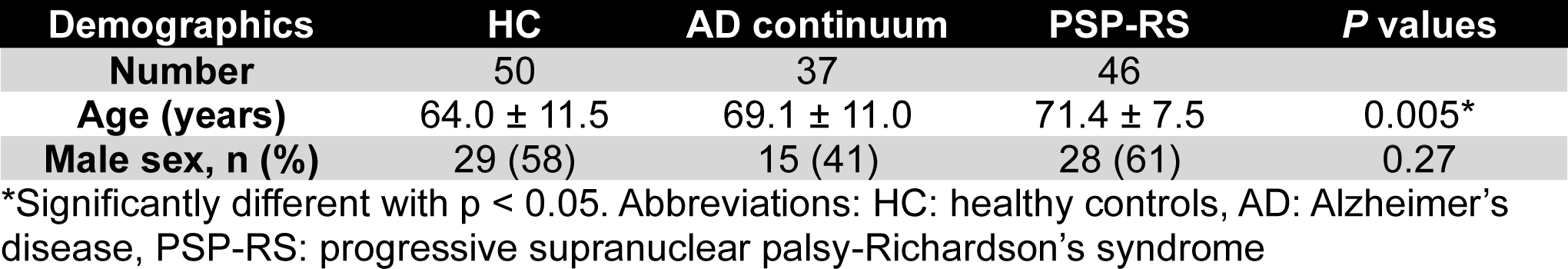
Demographic information.

| Demographics | HC | AD continuum | PSP-RS | *P* values |
| --- | --- | --- | --- | --- |
| **Number** | 50 | 37 | 46 |  |
| **Age (years)** | 64.0 ± 11.5 | 69.1 ± 11.0 | 71.4 ± 7.5 | 0.005* |
| **Male sex, n (%)** | 29 (58) | 15 (41) | 28 (61) | 0.27 |

\*Significantly different with $p < 0.05$ . Abbreviations: HC: healthy controls, AD: Alzheimer's disease, PSP-RS: progressive supranuclear palsy-Richardson's syndrome

TAME-Q processed all T1-weighted MR and florzolotau PET images successfully, with no failures.

In ROI-level analyses, the SUVR for the ITG was significantly higher in the AD group compared to both the HC and PSP groups (p < 0.001). In contrast, the SUVR for the GP was elevated in the PSP group relative to the HC and AD groups (p < 0.001) (Fig. 2).

**Fig. 2.**
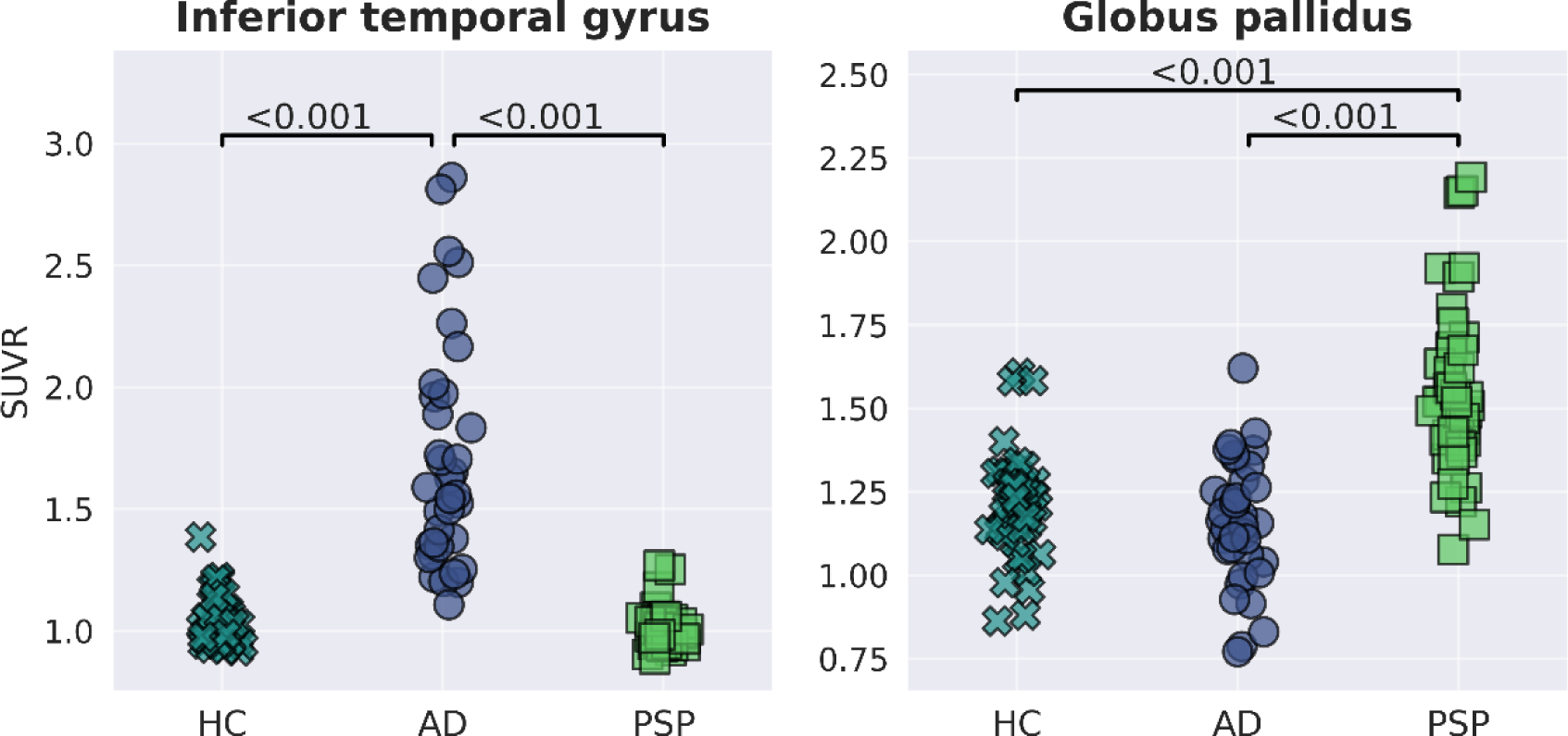
Group comparison of each ROI-level SUVR between groups. AD exhibited significantly higher SUVR in the inferior temporal gyrus compared to both HC and PSP. Conversely, PSP exhibited significantly elevated SUVR in the globus pallidus relative to both HC and AD. Group comparisons were conducted using Welch’s t-test.

Elastic Net training yielded mean hyperparameters (± SD) of *L1_ratio* = 0.641 ± 0.408 and *α* = 0.249 ± 0.223 for the AD-tau score, and *L1_ratio* = 0.365 ± 0.373 and *α* = 0.211 ± 0.141 for the PSP-tau score. As a result, 48 out of 133 ROIs had non-zero coefficients for AD-tau score, and 57 of 133 ROIs had non-zero coefficients for PSP-tau score (Supplementary Figure S1).

The AD-tau and PSP-tau scores showed significant correlations between the benchmark method and the proposed method, with correlation coefficients of 0.983 (95% CI: 0.976–0.988, p < 0.0001) for the AD-tau score and 0.966 (95% CI: 0.952–0.975, p < 0.0001) for the PSP-tau score.

The ROC analysis revealed an AUC of 0.999 (95% CI: 0.998–1.000) for the AD-tau score in discriminating AD and an AUC of 0.977 (95% CI: 0.959–0.997) for the PSP-tau score in discriminating PSP. DeLong’s test confirmed no significant differences in AUC between the benchmark and proposed methods for either score (AD-tau: Z = 0.820, p = 0.412; PSP-tau: Z = 0.570, p = 0.570) (Fig. 3).

**Fig. 3.**
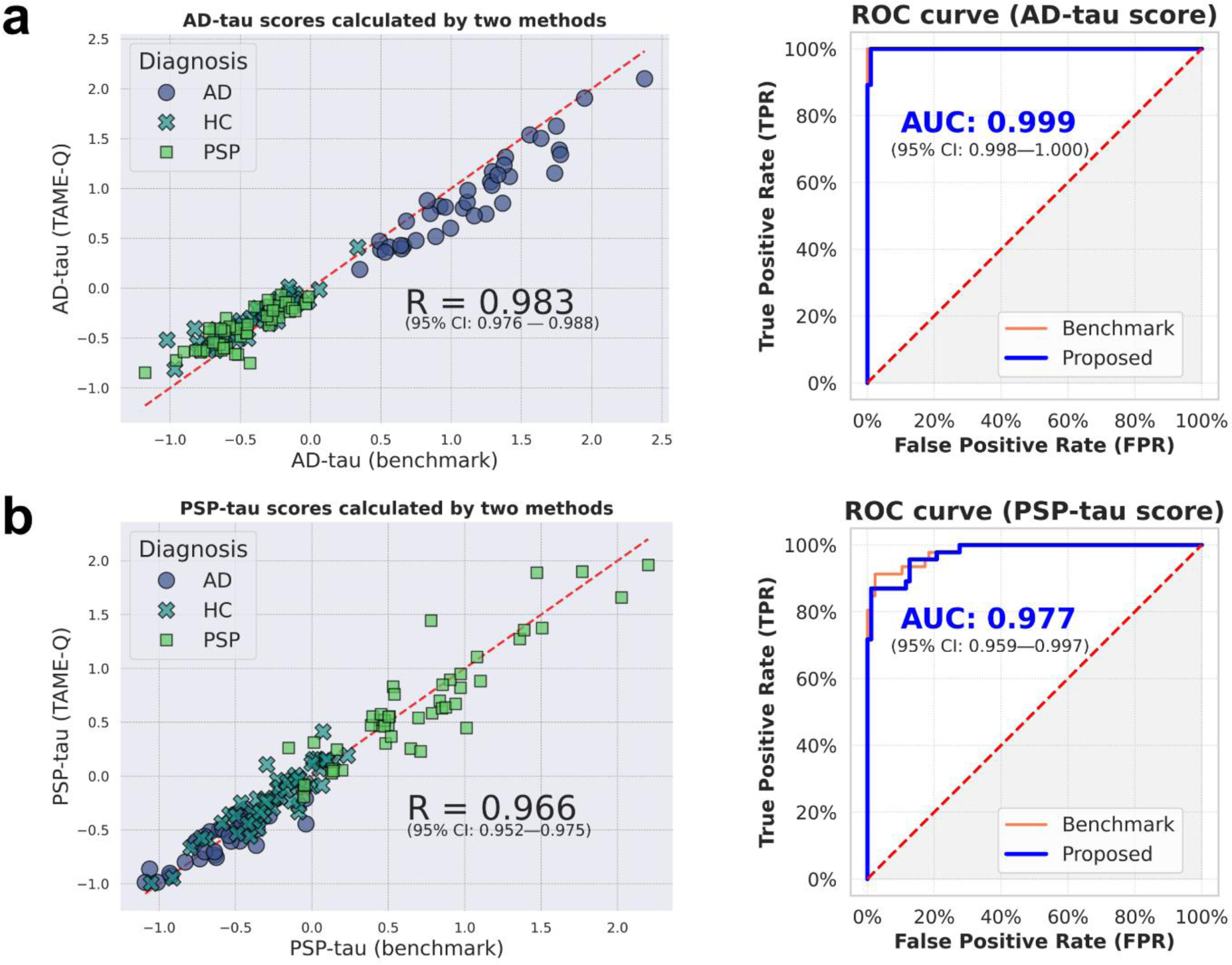
Correlation between tau scores across methods. (a) Scatter plots of AD-tau scores generated by the benchmark method and the proposed method, showing a correlation (R = 0.983, p<0.0001). ROC curves based on the AD-tau scores from each method are displayed. The area under the curve (AUC) for the proposed method was 0.999. A DeLong’s test revealed no statistically significant difference between the ROC curves of the benchmark and proposed methods. (b) Scatter plots of PSP-tau scores generated by the benchmark method and the proposed method, showing a correlation (R = 0.966, p<0.0001). ROC curves based on the PSP-tau scores from each method are displayed. The AUC for the proposed method was 0.977. A DeLong’s test revealed no statistically significant difference between the ROC curves of the benchmark and proposed methods.

## 4 Discussion

Here, we have presented TAME-Q, a comprehensive and automated preprocessing pipeline for dynamic PET images, incorporating grey-matter-based semi-quantification. In the present study, TAME-Q processed all target datasets without errors, and visual quality assurance did not identify any cases requiring manual correction. Consequently, the ROI-level SUVR results were obtained directly from the input data without any manual intervention. This minimal manual involvement reduces the burden of analysis and contributes to ensuring reproducibility. While visual evaluation for quality assurance is still necessary, the reports and histogram images simultaneously generated by TAME-Q facilitate this process. Also, alignment and curve-fitting parameters are served as table formats, allowing for the identification of erroneous cases by detecting outliers in these parameters when analyzing large datasets. Unlike earlier studies that relied on in-house scripts and proprietary software, TAME-Q is built entirely on open-source software and codes, thereby enhancing the transparency of preprocessing steps.

Furthermore, TAME-Q replicated known disease-specific SUVR elevations, thereby confirming the validity of its semiquantitative analysis. In brain regions corresponding to early Braak stages [42]—specifically the ITG—AD patients exhibited elevated SUVRs.

Likewise, PSP patients also demonstrated elevated SUVR in the GP, a region previously implicated in PSP studies as exhibiting increased tau signals [27]. Although we used a different parcellation scheme from earlier florzolotau investigations, our method captures comparable region-level SUVR elevations. This finding not only supports the validity of our preprocessing pipeline but also suggests that florzolotau permits atlas-independent quantitative analysis.

By integrating information from multiple ROIs, TAME-Q-derived SUVRs capture whole-brain tau patterns and avoid the instability of single-ROI classifiers (e.g., traumatic brain injury [43,44]). Elastic Net penalizes model complexity while retaining the most informative features, enabling the selective extraction of critical regions across the whole brain and their use in a quantitative evaluation. The Elastic Net model reconfigured in this study exhibited high discriminative performance, supporting the appropriateness of our processing pipeline. Moreover, the TAME-Q tau scores correlated strongly with scores produced in the benchmark study, despite differences in parcellation schemes and feature dimensionality. This high concordance confirms that our scores encode the same disease signatures identified in previous work.

These results demonstrate that the TAME-Q pipeline provides significant advantages for studying non-AD tauopathies. The ability of florzolotau to bind with all tau isoforms makes it a valuable tracer beyond AD, but only if semi-quantification is accurate. Traditional cerebellar referencing enables only group-level and visual assessments [45]. For objective stratification and quantitative assessment, methods that do not presuppose an intact cerebellum are more appropriate. By providing a histogram-based reference selection, TAME-Q facilitates objective, quantitative stratification across diverse tauopathies.

Beyond tau imaging, TAME-Q has potential for future applications with PET imaging modalities beyond tau PET. As diverse molecular imaging techniques continue to evolve, efforts are underway to visualize targets such as α-synuclein, TAR DNA-binding protein 43, and α-amino-3-hydroxy-5-methyl-4-isoxazolepropionic acid receptors using PET. In these contexts, the selection of an appropriate semi-quantification method constitutes a critical factor. The histogram-based approach implemented in TAME-Q, which does not require designating a single ROI as a reference, offers versatility across various scenarios. Consequently, TAME-Q enables straightforward access to histogram-based semi-quantification from the exploratory stage of these investigations.

A notable limitation of this study is that the data used were collected from a single institution. Although we did not include procedures that require specific conditions, using differing parameters (e.g., resolution, imaging devices) may give rise to unforeseen errors. As a current countermeasure, the pipeline produces reports and processing parameters so that any errors can be readily detected. These measures simplify quality checks and ensure the reliability of the preprocessing performance.

TAME-Q has been released as an open-source pipeline utility, and it is expected that, in the future, it will be applied under diverse conditions which will streamline multi-center collaborative research by ensuring consistent preprocessing across sites and foster broader advancements in imaging-based biomarker discovery.

## 5 Conclusions

TAME-Q processed semi-quantification of PET images accurately without manual intervention, and the processed images were useful for disease discrimination. By employing a histogram-based approach for preprocessing florzolotau PET images, TAME-Q is expected to enhance the reproducibility, transparency, and accessibility of the workflow, thereby facilitating multicenter validation. Released under a permissive license, it is readily deployable for multicenter studies and adaptable to other PET tracers. We expect that TAME-Q will accelerate biomarker validation and foster collaboration in imaging research across diverse tauopathies and molecular targets.

## Acknowledgements

The authors thank Yoko Ikoma, the engineer who developed the original code for the semi-quantification, for sharing it with us. We appreciate APRINOIA Therapeutics for generously providing a precursor and a reference standard of florzolotau (18F). This research was supported by JST SPRING, Grant Number JPMJSP2124.

## Statements and Declarations

### Ethics approval

This study was approved by the Radiation Drug Safety Committee and the National Institutes for Quantum Science and Technology Certified Review Board of Japan, and the Ethics Committee of the University of Tsukuba Hospital in accordance with the Declaration of Helsinki (R02-328).

### Consent to participate

Written informed consent was obtained from all participants. For participants with cognitive impairments, written informed consent was additionally obtained from their spouses or close relatives as appropriate.

### Funding

This work was funded in part by Japan Society for the Promotion of Science, KAKENHI, under Grant Number 25K15384; by AMED under Grant Numbers JP21dk0207055, JP23wm0625001, and JP24wm0625505; and JST under Grant Numbers JPMJCR1652 and JPMJMS2024.

### Competing Interests

M. Higuchi holds patents on compounds related to the present report (JP 5422782/EP 12 884 742.3/CA2894994/HK1208672). All other authors declare no conflict of interest.

### Author Contributions

All authors contributed to the study design. Material preparation and data collection were performed by Hironobu Endo, Kenji Tagai, Takahiko Tokuda, and Makoto Higuchi. Methodological implementation was performed by Kenjiro Nakayama and Kiyotaka Nemoto. Analysis was performed by Kenjiro Nakayama, Hironobu Endo, and Asaka Oyama. The first draft of the manuscript was written by Kenjiro Nakayama. All authors commented on previous versions of the manuscript. All authors read and approved the final manuscript.

### Data Availability

The data that support the findings of this study are available from the corresponding author upon reasonable request. The code for preprocessing images can be found in GitHub (https://github.com/kytk/tame-q).

---

**Supplementary Figure S1.**
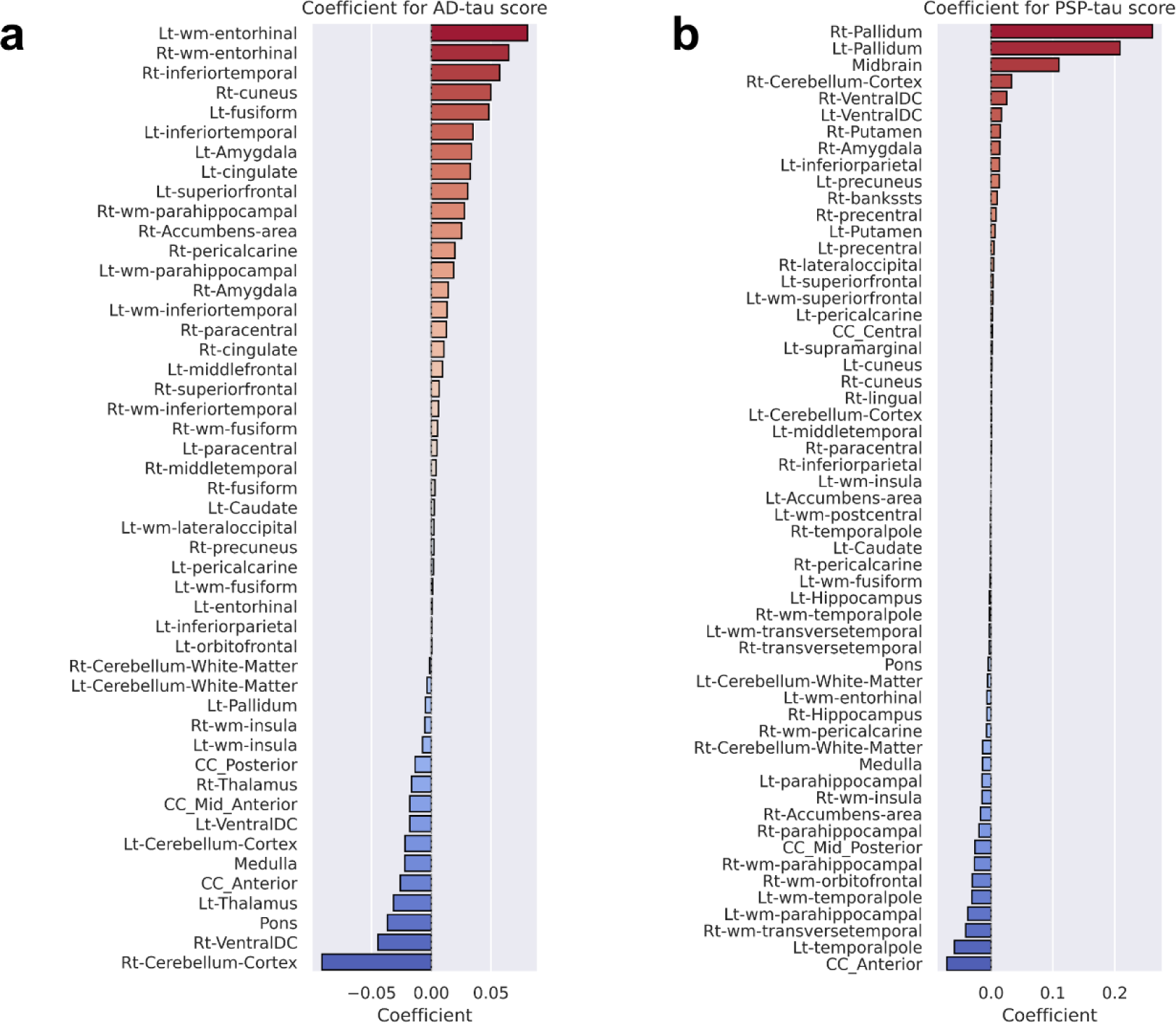
Coefficients for ROIs used to calculate tau scores obtained by the proposed method. (a) AD-tau scores had positive coefficients mainly in temporal lobes, including the entorhinal cortex and inferior temporal gyrus, and negative coefficients in the cerebellum. (b) PSP-tau scores had positive coefficients mainly in the globus pallidus and midbrain. ROI labels in the figure are primarily based on FreeSurfer ROI labels. A comprehensive list of their definitions is provided in Supplementary Table S1. Abbreviations: Lt = left; Rt = right; wm = white matter.

**Supplementary Table S1.**
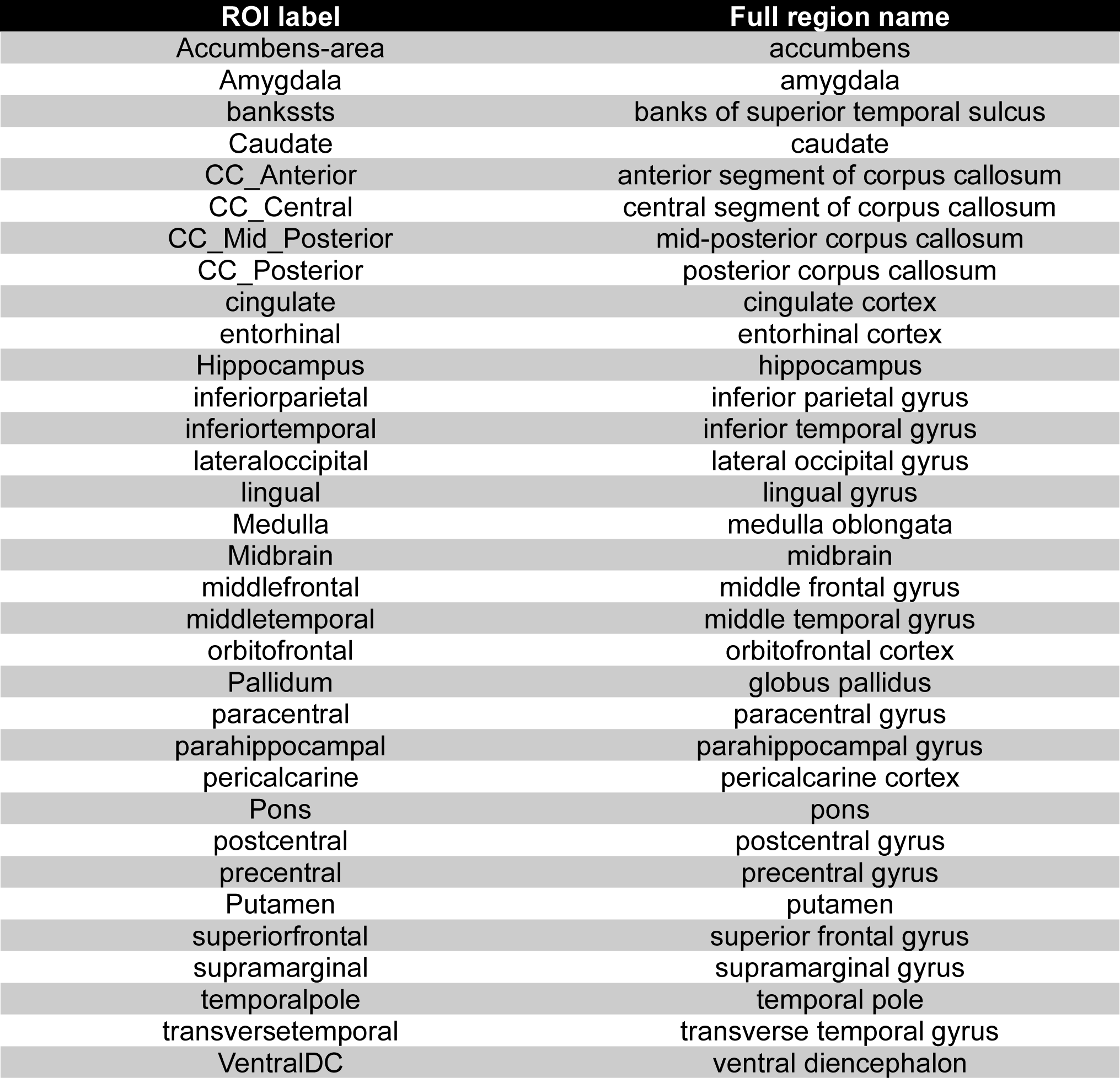
List of ROI labels.

## Notes

### Author Declarations

This study was approved by the Radiation Drug Safety Committee and the National Institutes for Quantum Science and Technology Certified Review Board of Japan, and the Ethics Committee of the University of Tsukuba Hospital in accordance with the Declaration of Helsinki.

